# Outcomes of endoscopic ultrasound-guided transmural drainage for postoperative peripancreatic fluid collection with an external drainage-first approach

**DOI:** 10.1101/2024.09.30.24314576

**Authors:** Jun Noda, Yuichi Takano, Naoki Tamai, Masataka Yamawaki, Tetsushi Azami, Fumitaka Niiya, Fumiya Nishimoto, Masatsugu Nagahama

## Abstract

**Objectives:** Endoscopic ultrasound-guided transmural drainage (EUS-TD) is widely performed to treat postoperative peripancreatic fluid collection (POPFC). Recent reports on EUS-TD lack a consensus on stent selection. This study aimed to assess the efficacy of EUS-TD for POPFC using an external drainage-first approach.

**Methods:** We retrospectively examined the medical records of patients with POPFC treated with EUS-TD using external drainage between October 2016 and July 2024. Technical success was defined as successful placement of the external drainage. Clinical success was defined as the disappearance of fluid collection on follow-up computed tomography without additional intervention.

**Results:** This study included 14 patients. The median duration from surgery to endoscopic treatment was 13 (range: 11–26) days. The median procedural time was 26 (range: 13–35) min. The technical success rate was 100%, and 6 Fr endoscopic nasocystic drainage was performed in all patients. The clinical success rate was 64.3%, and no adverse events were observed. In 35.7% (5/14) of the patients, additional endoscopic internal drainage was required; the POPFC was successfully controlled in all patients. The reasons for conversion to internal drainage were prolonged inflammation in one patient, remaining fluid collection in one patient, and requests from surgeons in three patients.

**Conclusions:** EUS-TD for POPFC with external drainage proved to be safe and effective, with a short procedure time. However, in certain patients, additional internal drainage is required.

## Introduction

Postoperative peripancreatic fluid collection (POPFC) primarily occurs because of pancreatic fluid leakage, leading to adverse events, such as infection, hemorrhage, and fistula formation. Pancreatic fluid leakage reportedly occurs in 5–20% of patients after pancreaticoduodenectomy (PD) [1] and 10–40% of patients after distal pancreatectomy (DP) [2]. While POPFC can be asymptomatic, it may manifest as intra-abdominal bleeding, severe pain, gastric outlet obstruction, fistula formation, intra-abdominal abscess, and sepsis [3,4].

Traditionally, POPFC has been managed via percutaneous or surgical drainage. In 2009, Varadarajulu first reported the application of endoscopic ultrasound-guided transmural drainage (EUS-TD) in 10 patients after DP, demonstrating high technical and clinical success rates with minimal adverse events [5]. Several studies recommend EUS-TD as the primary treatment method for POPFC management [6–9]. Regarding the optimal timing of EUS-TD intervention for POPFC, one meta-analysis identified six retrospective studies encompassing 128 and 107 patients who underwent early and delayed EUS-TD, respectively, from 1,415 initially screened articles [10]. They reported a drainage window ranging from 14 to 30 days, within which POPFC could be effectively managed using early EUS-TD without a high number of adverse events.

However, optimal drainage techniques, including appropriate stent selection and the duration of stent placement, remain unclear. Notably, literature focusing specifically on optimal drainage strategies for EUS-TD in the context of POPFC is sparse. In this study, we present our EUS-TD strategy for POPFC using an external drainage-first approach.

## Materials and Methods

### Patient selection

We retrospectively examined the medical records of patients with POPFC treated with EUS-TD using external drainage between October 2016 and July 2024 at Showa University Fujigaoka Hospital. The exclusion criteria were EUS-TD treatment using internal drainage only, both internal and external drainage, or percutaneous drainage, and reoperation.

### Definitions

POPFC was defined according to the guidelines established by the International Study Group for Pancreatic Surgery (ISGPS) on pancreatic drainage [11]. POPFC was defined as an abnormal communication between the pancreatic ductal epithelium and another epithelial surface containing pancreas-derived, enzyme-rich fluid. POPFC was also defined as fluid leakage of any measurable volume through an operatively-placed drain with an amylase activity greater than three times the upper limit of normal serum. In addition, a clinical system of three discrete grades of POPFC (grades A, B, and C) was proposed based on the severity of the complication. Technical success was defined as the successful placement of an external drain. Clinical success was defined as the disappearance of fluid collection on follow-up computed tomography (CT) without additional interventions. Adverse events were defined according to the criteria outlined by the American Society for Gastrointestinal Endoscopy. Procedural time was defined as the time from scope insertion to scope removal.

### EUS-TD strategy outline

Whether EUS-TD should be performed was determined through a comprehensive evaluation by surgeons and endoscopists based on follow-up CT, percutaneous drain output, amylase levels in the drainage fluid, patient symptoms (abdominal pain and fever), and serum C-reactive protein (CRP) levels, leading to a consensus.

For patients in whom POPFC was confirmed, the steps of the EUS-TD procedure were as follows (Fig 1): When the patient had been placed under sedation with 35 mg of pethidine hydrochloride and 2–5 mg of midazolam, an Olympus convex array endoscope (GF-UCT260; Olympus Medical Systems, Tokyo, Japan) was inserted (Fig 1B). Peripancreatic fluid collection was confirmed from the gastric lumen, and Doppler-guided puncture was performed at a site devoid of intervening vessels. The puncture was performed using a 19 G needle (EZ shot3, Olympus), and a 0.025-inch guidewire (Visiglide2, Olympus) was inserted into the fluid cavity. After dilation using a 7 Fr mechanical dilator (ES dilator, Zeon Medical, Tokyo, Japan) or balloon catheter (REN, Kaneka, Yokohama, Japan), a 6 Fr pigtail-type endoscopic nasocystic drain (ENCD) (Cook Medical) was inserted in all patients (Fig 1C).

**Fig 1.**
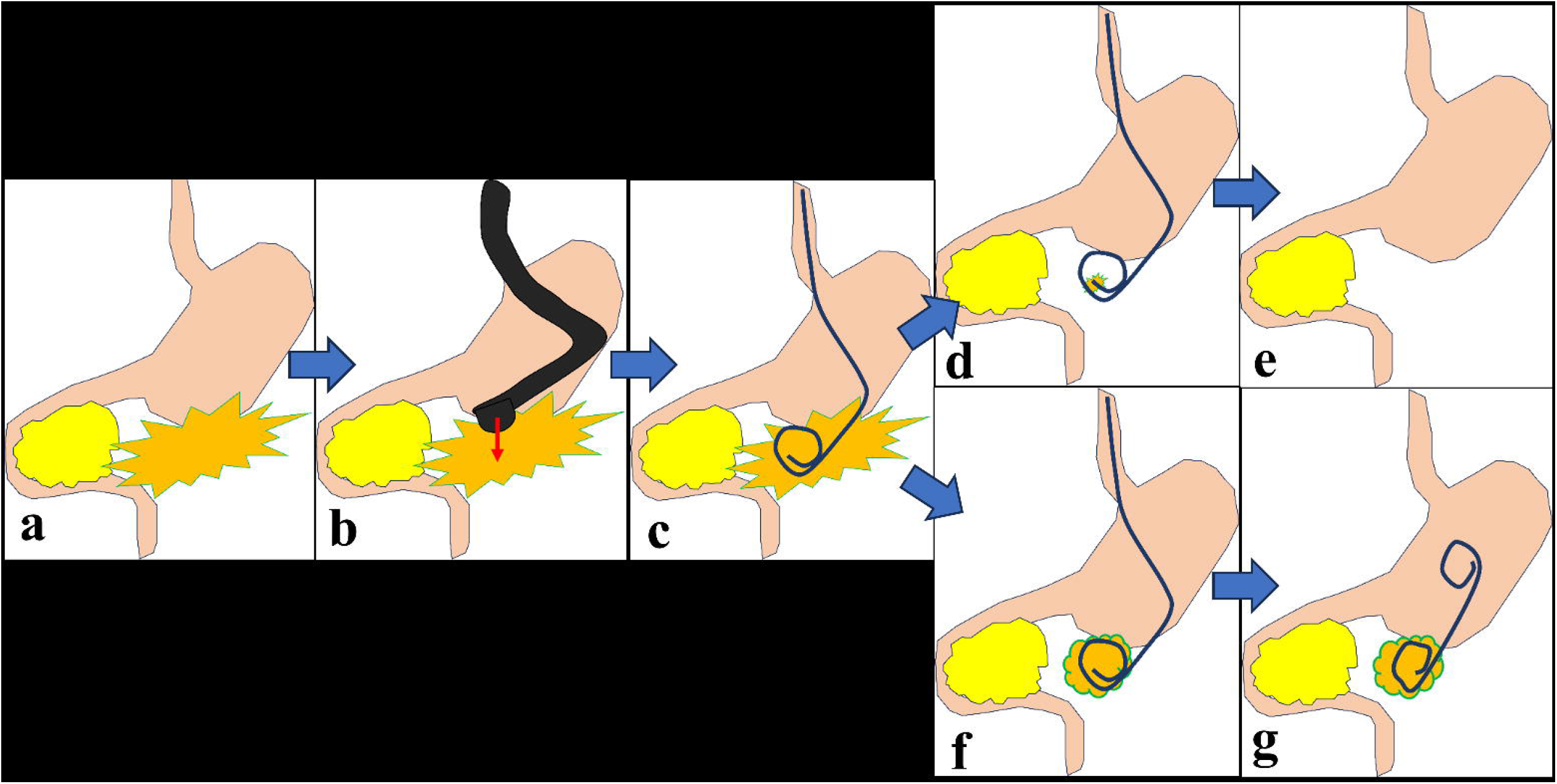
Study flow diagram. POPFC, postoperative peripancreatic fluid collection; EUS-TC, endoscopic ultrasound-guided transmural drainage

The timing of external drain removal was based on the disappearance of clinical symptoms, such as fever and abdominal pain, improvement in serum CRP levels, and disappearance of fluid collection on follow-up CT 1 week later. When the cavity had disappeared (Fig 1D), the external drain was removed (Fig– 1E). In cases of poor improvement or upon request from the surgeons (Fig 1F), internal drain replacement was planned. First, an attempt was made to insert a guidewire into the cyst through the side of the external drain. It was often necessary to re-puncture as the fluid volume shrank, and a contrast agent was injected through the external drain to expand the area. Finally, internal drainage was performed to complete the procedure (Fig 1G). A 7 Fr double-pigtail-type stent (Zimmon, Cook, or Through & Pass; Gadelius Medical, Tokyo, Japan) was used; the stent was left in place permanently for the duration of outpatient care.

### Ethical statement

This retrospective study was reviewed and approved by the Ethics Committee of Showa University (approval no. 21-061-A). This study was conducted in accordance with Good Clinical Practice principles in accordance with the Declaration of Helsinki. Data were accessed for research purposes from December 1, 2019 to May 31, 2024.The requirement for informed consent was waived due to the retrospective nature of the study and utilization of anonymous data.

## Results

### Patient characteristics

A retrospective cohort of 17 patients who underwent EUS-TD for POPFC at Showa University Fujigaoka Hospital between October 2016 and July 2024 was included. Of these, three patients, in whom internal and external drainage were selected, were excluded (Fig 2). Finally, 14 patients who underwent the external drainage-first approach were included; their characteristics are presented in Table 1. The median age was 74.5 (range: 51–84) years, and the group included 10 men and 4 women. The most prevalent primary disease was pancreatic cancer, which was observed in nine patients. The surgical procedures included DP in 11 patients, PD in 2 patients, and combined PD with DP in 1 patient. All patients were categorized as grade B, according to the ISGPS classification system.

**Fig 2.**
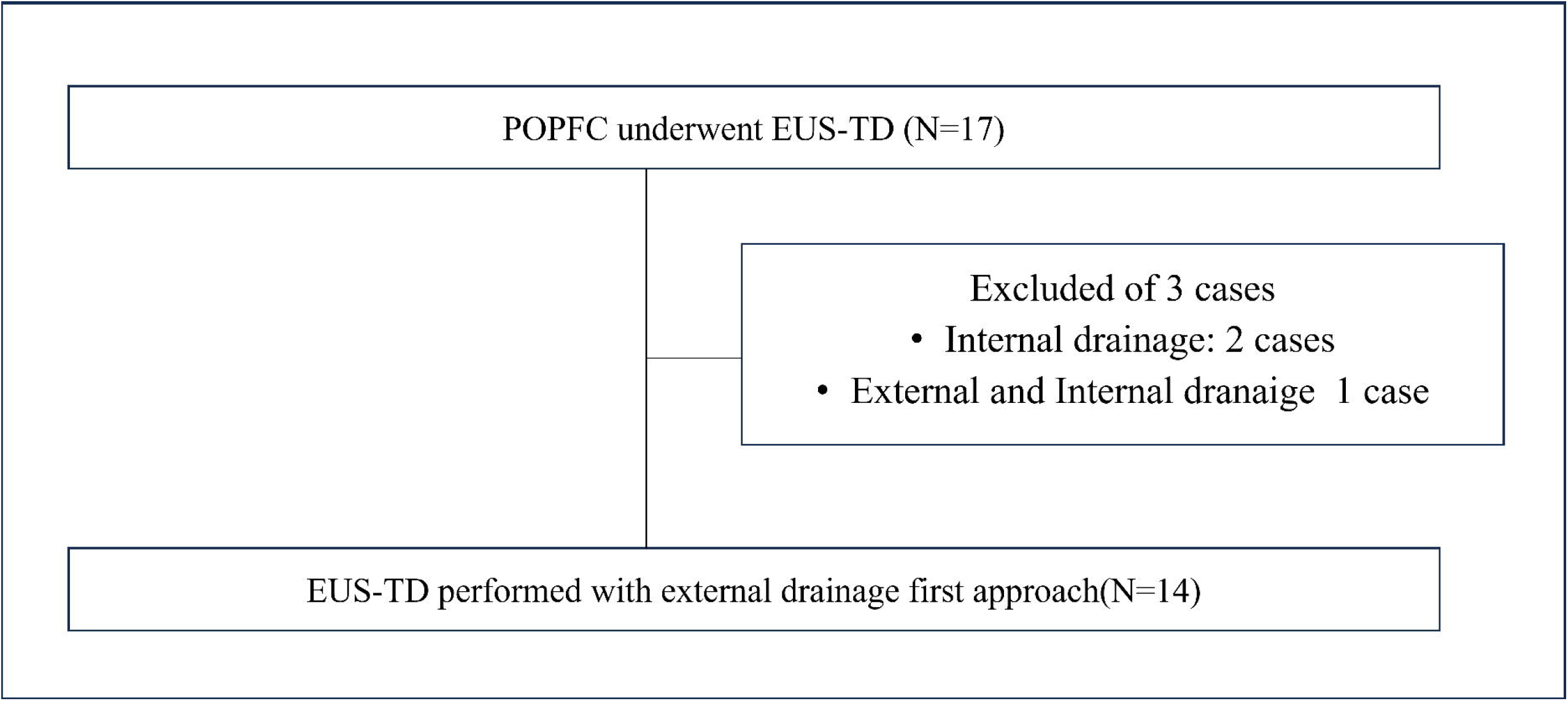
(A) POPFC was confirmed, and (B) EUS-TD was performed. (C) We inserted the external drain only. (D) Once the cavity had disappeared, (E) the external drain was removed. (F) If the cavity persisted, (G) external drainage was changed to internal drainage. POPFC, postoperative peripancreatic fluid collection; EUS-TD, endoscopic ultrasound-guided transmural drainage.

**Table 1:** Characteristics of patients who underwent EUS-TD for POPFC.

| Table 1: Characteristics of patients who underwent EUS-TD for POPFC. |  |  |  |  |
| --- | --- | --- | --- | --- |
| Case | Age | Sex | Disease | Surgical Method |
| 1 | 61-65 | M | PDC | PD + DP |
| 2 | 51-55 | M | PDC | DP |
| 3 | 71-75 | F | PDC | PD |
| 4 | 81-85 | M | BTC | PD |
| 5 | 81-85 | M | AIP | DP |
| 6 | 71-75 | M | IPMN | DP |
| 7 | 81-85 | M | PDC | DP |
| 8 | 81-85 | M | PDC | DP |
| 9 | 51-55 | M | PDC | DP |
| 10 | 76-80 | M | PDC | DP |
| 11 | 65-70 | F | IPMN | DP |
| 12 | 51-55 | M | PNET | DP |
| 13 | 76-80 | F | PDC | DP |
| 14 | 81-85 | F | PDC | DP |

EUS-TD, endoscopic ultrasound-guided transmural drainage; POPFC, postoperative pancreatic collection; PDC, pancreatic duct cancer; IPMN, intraductal papillary mucinous neoplasm; PNET, pancreatic neuroendocrine tumor; BTC, Biliary tract cancer; AIP, Autoimmune pancreatitis; PD, pancreatoduodenectomy; DP, distal pancreatectomy;

### Outcome of EUS-TD for POPFC

The EUS-TD outcomes are shown in Table 2. The median duration from surgery to endoscopic treatment was 13 (range: 11–26) days, with a median procedural time of 26 (range: 13–35) min. The median duration of the removal of the external drainage was 11.5 (range: 7–23) days, and the median postprocedural hospital stay was 16 (range: 10–32) days.

**Table 2:**
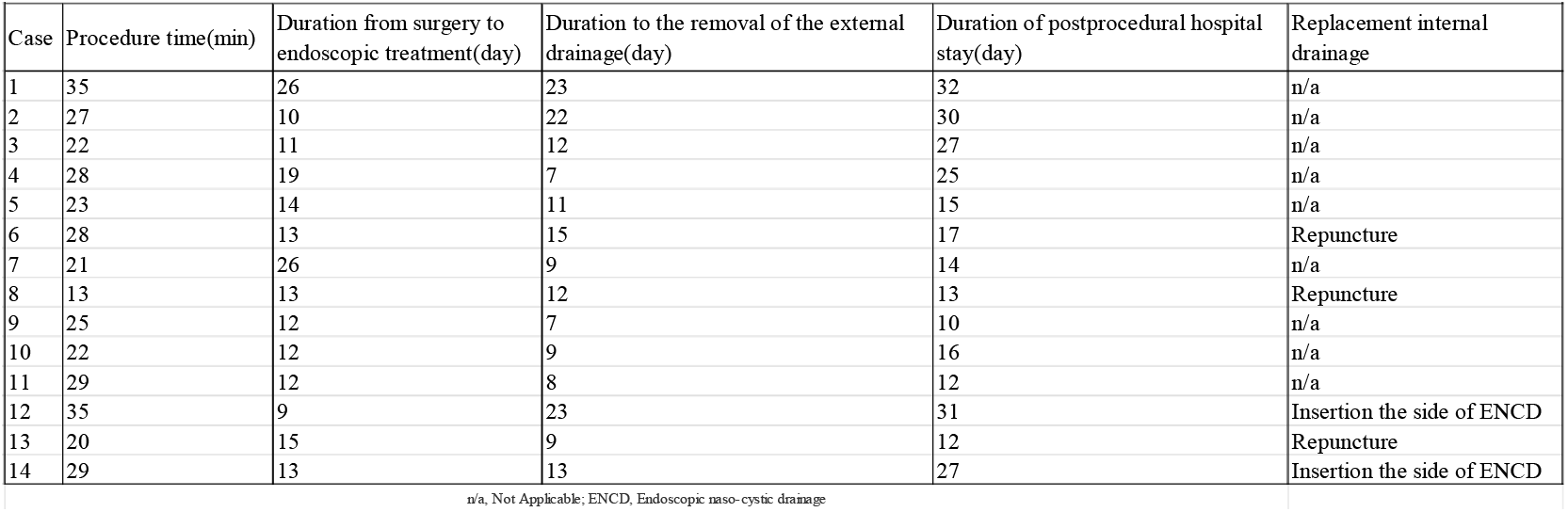
Details of endoscopic treatment.

The technical success rate was 100%, and the clinical success rate was 64.3%, with no occurrences of adverse events, such as bleeding or perforation. Self-removal occurred in one patient, and a second EUS-TD was performed.

Regarding progress after stent placement, nine (64.3%) patients had no recurrence after removal of the external drain, while five (35.7%) patients required additional EUS-TD for internal drainage. In two patients, the internal drainage tube was successfully inserted beside the external drainage tube using forward-viewing gastroduodenoscopy (GIF-Q260J, Olympus). In three patients, a second EUS-TD was required. The reasons for conversion to internal drainage were prolonged inflammation in one patient, remaining fluid collection in one patient, and requests from surgeons in three patients.

Table 3 shows the changes in CRP and amylase levels in the drainage fluid, and the culture results of the treated patients. In all tested patients, amylase levels were very high and the cultures were positive. In some patients, CRP levels temporarily increased after treatment but improved after 7 days.

**Table 3:** Details of endoscopic treatment.

| Table 3: Details of endoscopic treatment. |  |  |  |  |  |
| --- | --- | --- | --- | --- | --- |
| Case | Before treatment<br>CRP levels (mg/dl) | Day3 after treatment CRP levels (mg/dl) | Day7 after treatment<br>CRP levels (mg/dl) | Amy levels of<br>drainage(U/L) | Culture of drainage |
| 1 | 3.44 | 6.09 | 4.6 | n/a | Intestinal bacteria |
| 2 | 3.22 | 1.88 | 1.41 | 3058 | n/a |
| 3 | 24.95 | 12.86 | 4.38 | 10018 | Fungus |
| 4 | 23.77 | 15.81 | 7.27 | 15825 | *Enterococcus.* |
| 5 | 4.33 | 3.97 | 1 | 5666 | Intestinal bacteria |
| 6 | 11.66 | 4.84 | 4.4 | 2774 | Intestinal bacteria |
| 7 | 2.06 | 6.35 | 0.62 | 36404 | *Enterococcus.* |
| 8 | 2.48 | 4.12 | 3.23 | 24900 | Intestinal bacteria |
| 9 | 7 | 2.46 | 0.64 | 50611 | *Enterococcus.* |
| 10 | 19.8 | 2.42 | 0.49 | 37810 | *Enterococcus.* |
| 11 | 1.44 | 3.83 | 0.28 | 1714 | *Streptococcus.* |
| 12 | 5.91 | 2.76 | 1.47 | 35748 | n/a |
| 13 | 2.34 | 1.21 | 0.34 | 103555 | *Staphylococcus.* |
| 14 | 15.43 | 0.91 | 0.37 | 21167 | *Enterococcus.* |
| CRP, C-reactive protein; Amy, Amylase; n/a, Not Applicable. |  |  |  |  |  |

### Case presentations

Case 1: A woman in her 70s with a history of PD for cancer of the pancreatic head underwent EUS-TD on the 11^th^ postoperative day. CT revealed peripancreatic fluid collection around the pancreaticojejunostomy site (Fig 3A), and EUS revealed amorphous fluid collection around the pancreas (Fig 3B). Using a 19 G needle, puncture was performed from the gastric body, and a guidewire was placed at the site of fluid collection. After dilation using an ES dilator, a 6 Fr pigtail-type ENCD was inserted. Drainage contrast imaging confirmed adequate placement of the ENCD at the peripancreatic fluid collection site (Fig 4A), and the procedure was concluded. On postoperative day 3, CRP levels improved from 24.95 to 4.95 mg/dL on the day after EUS-TD. Follow-up CT 1 week after drainage showed complete disappearance of the fluid collection (Fig 4B). Consequently, the ENCD was removed 13 days after EUS-TD, and the patient was discharged without recurrence.

**Fig 3.**
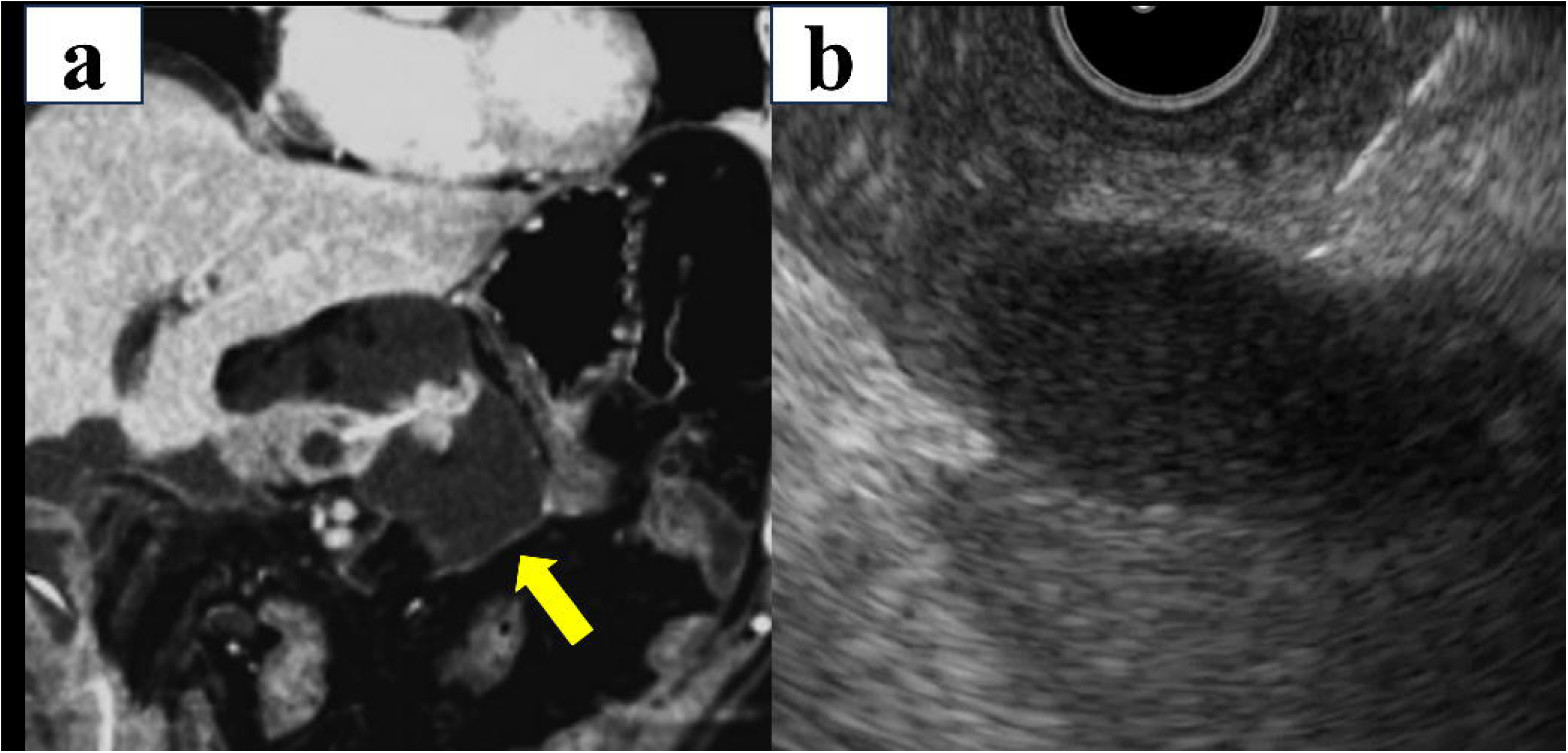
(A) CT performed 11 days after surgery shows POPFC (arrow). (B) EUS imaging shows POPFC, into which a 19 G needle was inserted trans-gastrically. CT, computed tomography; EUS, endoscopic ultrasound; POPFC, postoperative peripancreatic fluid collection

**Fig 4.**
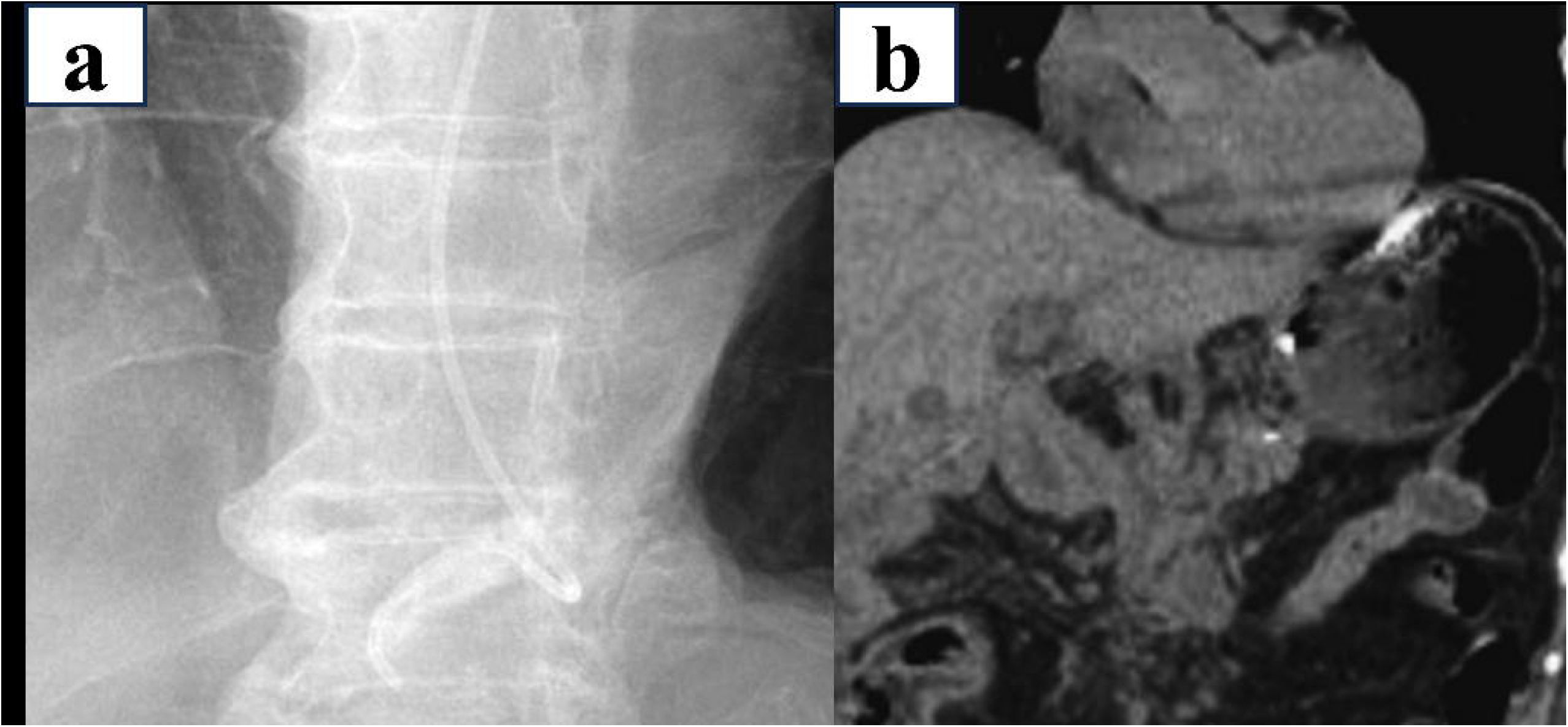
(A) Endoscopic ultrasound-guided drainage was performed, and a 6 Fr endoscopic nasocystic drainage tube was placed. (B) CT performed 7 days after EUS-TD showed that the fluid collection had completely disappeared. CT, computed tomography; EUS-TD, endoscopic ultrasound-guided transmural drainage. EUS, endoscopic ultrasound

Case 2: An man in his 70s had a history of DP for pancreatic cancer. CT findings 14 days after the surgery revealed peripancreatic fluid collection. A puncture was performed using a 19 G needle from the gastric body (Fig 5A), and a guidewire was placed at the site of fluid collection. Following this, the site was dilated using an ES dilator, after which a 6 Fr pigtail-type ENCD was inserted (Fig 5B). Follow-up CT conducted 27 days after surgery indicated a reduction in the size of the POPFC, although it remained present (Fig 6A); therefore, additional drainage was deemed necessary. The insertion of an internal drainage tube was planned; however, insertion from the side of the ENCD was difficult. On EUS, the POPFC was unclear. We attempted to expand the cavity by injecting a contrast medium through the ENCD. We were then able to perform a puncture with a 19 G needle, and a 0.025-inch guidewire was placed (Fig 6B). After dilatation with a 7 Fr mechanical dilator, a 7 Fr 7 cm double-pigtail-type stent was successfully placed (Fig 6C). No adverse events were observed after removal of the ENCD.

**Fig 5.**
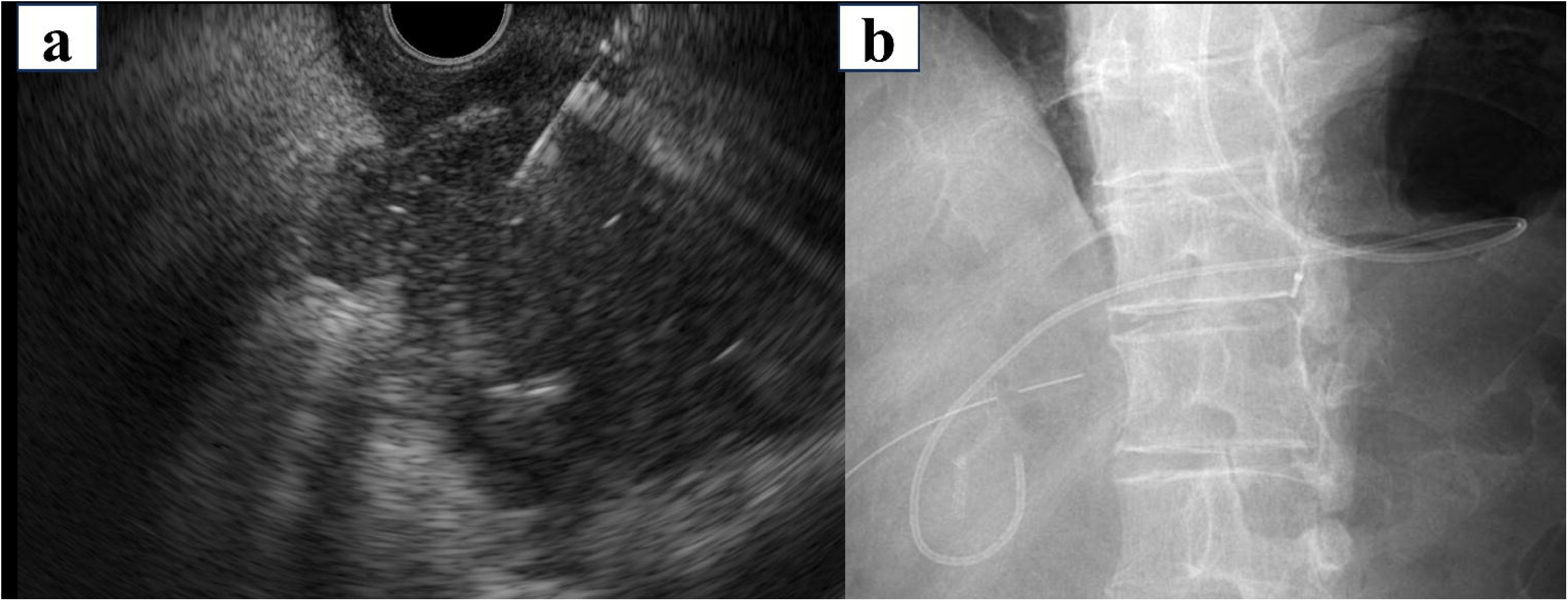
(A) EUS imaging shows POPFC, into which a 19 G needle was inserted trans-gastrically. (B) A 6 Fr endoscopic nasocystic drainage was successfully placed. EUS, endoscopic ultrasound; POPFC, postoperative peripancreatic fluid collection

**Fig 6.**
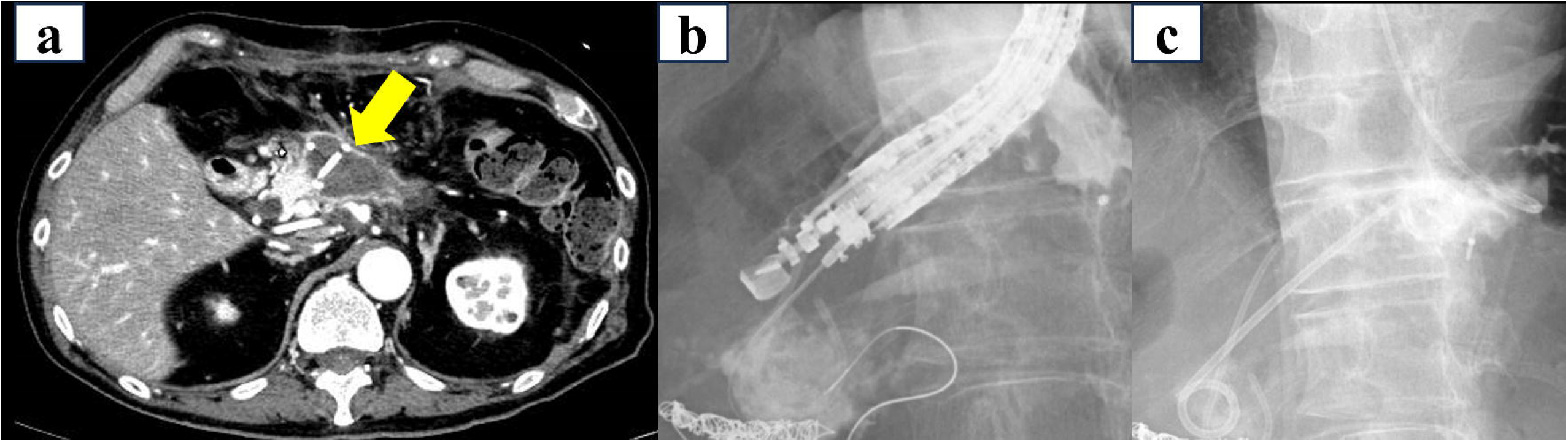
(A) Contrast-enhanced CT on the 27^th^ day after surgery showed a reduction in the size of the peripancreatic fluid collection; however, it remained present (arrow). (B) We were then able to perform a puncture with a 19 G needle, and a 0.025-inch guidewire was placed. (C) A 7 Fr 7 cm double-pigtail stent was successfully placed.

## Discussion

We retrospectively analyzed the data of 14 patients who underwent EUS-TD at our institution using the external drainage-first approach for POPFC between 2016 and 2024. We examined the technical and clinical outcomes and adverse events. The technical success rate was 100%, and EUS-TD for POPFC was considered a safe procedure. Previous studies have reported varied findings regarding the efficacy of EUS-TD for POPFC [6–9]. These studies have reported favorable outcomes, with technical and clinical success rates ranging from 92% to 100%. Moreover, the incidence of adverse events reportedly ranges between 0% and 8%, indicating favorable overall outcomes. Regarding the timing of EUS-TD, a meta-analysis reported that early intervention within 1 month was effective [10]. Our study also showed favorable results when EUS-TD was performed as early as 13 days after surgery. At our institution, EUS-TD for POPFC has been performed since 2016. When we first started performing EUS-TD, consultations with the surgeon were often conducted approximately 1 month postoperatively. However, requests for EUS-TD have recently been received as early as 10 days postoperatively. Early EUS-TD may be considered in patients with inflammation or infection.

No consensus has been reached on the appropriate stent selection for POPFC. At our institution, external drainage placement is generally selected during EUS-TD for POPFC. We were concerned about the risk of stent migration due to insufficient capsule formation in the early postoperative period. External drainage placement is useful for avoiding stent migration, measuring drainage output, and technical ease [8,10,12]. However, in one patient in our study, self-removal of the external drain led to the need for reintervention; internal drainage should be prioritized in patients with dementia or delirium. No consensus has been reached regarding the timing of external drain removal. Wang et al. reported waiting for encapsulation for 1 month while monitoring drainage and symptoms; if the cyst diameter was <2 cm, external drain removal was considered [12]. Leaving an external drain in place for too long can be uncomfortable for the patient. Our criteria for removal were symptom improvement and the disappearance of fluid collection on CT.

In this study, approximately 30% of the patients required additional drainage, and in some, re-puncture using an echoendoscope was required. If a cavity persists on follow-up CT or inflammation remains high, residual pus may be present, which cannot be drained using external drainage [7].

Some surgeons believe that internal and external drainage should be selected at the outset; however, we selected external drainage to avoid stent migration and for technical ease. Although the ENCD being cut for internal drainage has been reported [8], the remaining tube on the gastric side may straighten, raising concerns about migration. In addition, the gastric side of the tube has no side holes, which may result in poor drainage. We believe that pigtail-type internal drainage is preferable for the prevention of stent migration and the provision of sufficient drainage, considering that the stent will be placed permanently. It is often difficult to insert an internal drain because the cavity has already shrunk. As in Case 2, expansion through the external drain may be useful.

Large-diameter lumen-apposing metal stents (LAMS), such as the Hot AXIOS™ System (Boston Scientific Japan, Tokyo, Japan), have recently been used to treat post-pancreatitis pseudocyst and walled-off necrosis, aiming for effective drainage [13]. Overseas reports on drainage using LAMS to treat POPFC have shown favorable results [14]. The greatest advantage of LAMS is its superior drainage to that of plastic stents, owing to its larger diameter. For POPFC persisting for >1 month, fluid collection may already be encapsulated, and the risk of stent migration may be lower. However, after drainage, metal stents may come into contact with other organs in the abdominal cavity, leading to inflammation or bleeding (e.g., pseudoaneurysm) [15]. Additionally, LAMS is an expensive device, and its use should be carefully considered in terms of cost-effectiveness.

This study had several limitations. First, it was a retrospective study conducted at a single institution with a small number of patients. This retrospective design may have introduced biases that affected data collection and interpretation. Nevertheless, this study is the first to investigate optimal stent selection for EUS-TD for POPFC; it contributes valuable insights despite the aforementioned limitations.

## Conclusions

EUS-TD for POPFC with an external drainage-first approach proved to be safe and effective with a short procedure time. However, additional internal drainage is required in certain cases.

## Data Availability

All relevant data are within the manuscript and its Supporting Information files.

## Acknowledgments

JN would like to thank members of the Department of General and Gastroenterological Surgery, Showa University Fujigaoka Hospital, for their insightful discussions.

## Author contributions

JN, TY, TA, MY, and FN acquired patient data, and JN prepared the manuscript and figures. NT, FN, and MN participated in data acquisition and analysis. All authors agree with the contents of this manuscript.

